# A Bayesian hierarchical model with integrated covariate selection and misclassification matrices to estimate neonatal and child causes of death

**DOI:** 10.1101/2021.02.10.21251488

**Authors:** Amy R. Mulick, Shefali Oza, David Prieto-Merino, Francisco Villavicencio, Simon Cousens, Jamie Perin

## Abstract

Reducing neonatal and child mortality is a global priority. In countries without comprehensive vital registration data to inform policy and planning, statistical modelling is used to estimate the distribution of key causes of death. This modelling presents challenges given that the input data are few, noisy, often not nationally representative of the country from which they are derived, and often do not report separately on all of the key causes. As more nationally representative data come to be available, it becomes possible to produce country estimates that go beyond fixed-effects models with national-level covariates by incorporating country-specific random effects. However, the existing frequentist multinomial model is limited by convergence problems when adding random effects, and had not incorporated a covariate selection procedure simultaneously over all causes. We report here on the translation of a fixed effects, frequentist model into a Bayesian framework to address these problems, incorporating a misclassification matrix with the potential to correct for mis-reported as well as unreported causes. We apply the new method and compare the model parameters and predicted distributions of eight key causes of death with those based on the previous, frequentist model.

## Introduction

Reducing child (under-five) mortality has been a priority for individual countries and the broader international community for decades, but was given added impetus by the Millennium Development Goals (MDGs) established by the United Nations (UN) in 2000. Although dramatic reductions occurred over the MDG period until 2015, an estimated 5.3 million deaths still occurred worldwide in children under five in 2019 (UNICEF et al., 2019). Neonatal deaths (those in the first 28 days of life) account for 47% of under-5 child deaths (UNICEF et al., 2019), and have reduced more slowly than those in the 1–59 month age group (Hug et al., 2019). The Sustainable Development Goals (SDGs; 2015–2030), which followed on from the MDGs, include targets of 25 or fewer under-5 deaths and 12 or fewer neonatal deaths per 1,000 live births for all countries by 2030 (United Nations, 2017).

Understanding the cause-of-death (COD) distribution of neonatal and child deaths is important for selecting appropriate interventions to reduce mortality. Ideally, this information would be available through regularly updated, high-quality data collection systems that are organized at the national level but operate at the local level. At present, only around 70 countries have high-quality vital registration (VR) systems that regularly collect and collate causes of death (World Health Organization, 2017), and the majority of these are high-income countries with low mortality rates. Thus, statistical modelling remains an important tool for estimating the distribution of causes that lead to death for all age groups, including neonates.

Nationally comparable estimates of the neonatal and 1–59 month COD distributions have been produced for 190+ countries since 2005 by the Child Health Epidemiology Reference Group (CHERG) and Maternal Child Epidemiology Estimation (MCEE) group in collaboration with the World Health Organization (WHO). For countries without high-quality VR data, these distributions have been estimated using regression models within a frequentist framework (Liu et al., 2016). This existing approach has several strengths but some important limitations, including the approach to covariate selection, the lack of a mechanism to give additional weight to country-specific data for a country’s own estimates and the inability to account for misclassification in the reported CODs.

In this paper, we present our work on translating the existing COD estimation method into the Bayesian framework, extending it to include country-specific random effects and handle COD misclassification, and illustrate its application to neonatal deaths. In Sect. 1 we briefly describe the existing frequentist strategy and outline its limitations; in Sect. 2 we propose methods to address these limitations; in Sect. 3 we outline the model building process; in Sect. 4 we present and evaluate our results for neonatal deaths and compare them with results from the previous method; and we outline the strengths, limitations, and implications of this new modelling approach in the Discussion.

## 1. Existing strategy and limitations

To produce nationally comparable neonatal COD distributions, we classify each of 194 WHO member states (countries) into one of three groups: those with 1) high-quality VR data, 2) inadequate VR data and low child mortality rates, and 3) inadequate VR data and high child mortality rates (World Health Organization, 2018). For countries in the first group, we use their VR data directly to estimate proportional COD distributions. For countries with inadequate VR, we predict their COD proportions using a “low mortality model”(group 2 countries) or a “high mortality model” (group 3 countries). In this paper, we focus on the high mortality model as this model is more technically challenging and requires methodological innovations.

### 1.1 Data inputs

#### Cause-of-death data

For the high mortality model, we extracted relevant neonatal COD distributions from studies conducted in high mortality settings since 1980. Details of the literature review are found in (Oza et al., 2015). From the review, we identified 95 studies that reported causes for 100,119 neonatal deaths in 37 countries (range 1-18 studies per country) between 1980 and 2013. Twenty countries produced one study; only two (Bangladesh and India) produced more than 10.

These studies typically used verbal autopsy (VA) methods to ascertain the cause of death. This involves interviewing relatives about the symptoms experienced by the deceased individual and using this information to assign a cause of death (World Health Organization, 2016). These studies ranged widely in size and specific study methodologies. Where necessary, we re-classified the recorded CODs into eight key neonatal COD categories: 1) complications of preterm birth (“preterm”), 2) intrapartum-related complications (“intrapartum”), 3) congenital disorders (“congenital”), 4) sepsis and other severe infections (“sepsis”), 5) pneumonia, 6) diarrhoea, 7) neonatal tetanus (“tetanus”), and 8) other causes (“other”). Case definitions are detailed elsewhere (Oza et al., 2015).

#### Covariate data

We considered 14 explanatory variables for inclusion in our model. Nine of these were continuous metrics: under-five mortality rate (U5MR), neonatal mortality rate (NMR), general fertility rate (GFR), low birthweight rate (LBW), proportion of women delivering with a skilled birth attendant (SBA), adult female literacy rate (FLR), proportion of babies protected at birth against tetanus (PAB), diphtheria/pertussis/tetanus vaccine coverage (DPT), and Bacillus Calmette-Guerin vaccine coverage (BCG). The other five covariates were binary (yes/no) and relate to individual studies: whether reported deaths were from the early neonatal period (“per.early”; 0–6 days, reference 0–27 days); whether reported deaths were from the late neonatal period (“per.late”; 7–27 days, reference 0–27 days); whether the study was conducted in Sub-Saharan Africa (SSA); whether the study was conducted in South Asia (SA); and whether the study distinguished between prematurity and low birth weight (“premvslbw”).

### 1.2. Frequentist modelling approach

We selected “intrapartum” as our “baseline” cause for a multinomial model as all studies reported deaths due to intrapartum-related complications and they represent a relatively high proportion of deaths in high mortality settings. Our frequentist COD modelling approach followed three steps: 1) select covariates using logistic regression for each (non-baseline) COD equation; 2) with selected covariates, build a multinomial regression model for all causes simultaneously and obtain estimated model coefficients; and 3) apply the estimated model coefficients to national-level covariates to produce country-level COD distributions. The outcome for each regression equation in step 1 was the log of the ratio of deaths attributed to the given cause relative to deaths attributed to the baseline cause, and we used out-of-sample goodness of fit (GOF) under a jackknife (leave-one-out) procedure to select covariates for each equation.

Not all studies reported CODs in the eight key categories we model. To account for unreported causes, we re-wrote the multinomial likelihood function based on assumptions about which cause category deaths from an unreported cause would have been assigned. For example, if preterm, congenital, or sepsis were unreported, we assumed deaths from these to be in the “other” category. If pneumonia, diarrhoea, or tetanus were unreported, these were assumed to have been included in the sepsis/severe infection category.

Detailed methods have been described by Oza et al. (2015) and are summarised in **Online Supplement Text E1**.

### 1.3. Limitations of this modelling approach

These multinomial models have been used for fifteen years by the CHERG-MCEE-IMPROVE team, with various minor extensions and modifications over time, to produce neonatal COD estimates for the UN. However, some key issues led us to investigate further improvements and alternative modelling approaches.

First, our current modelling strategy does not give additional weight to input data from a given country for that country’s modelled estimates. Therefore, empirical data from a particular country do not influence that country’s modelled COD proportions any more than data from other countries. Previously, almost all the studies in our input database were small and not nationally representative, and it was not obvious that the national-level estimates for a country should be particularly influenced by data points from small non-national studies. However, an increasing number of countries now have data from nationally representative VA studies, and efforts are underway to increase the number of such studies (COMSA, 2020).

Second, the current covariate selection approach is an efficient method to search over a large space of covariate combinations. However, two potential limitations with this method are that 1) it does not evaluate all possible covariate combinations and 2) we select covariates for a multinomial model using binomial models. We used individual binomial equations for covariate selection because a similar multinomial approach would be computationally prohibitive.

Finally, apart from the out-of-sample approach to covariate selection, there are no other stability-enhancing components in the modelling process to minimize the impact of noisy data and the risk of overfitting. The input data in our models contains substantial noise due to measurement error in both the outcome (COD) and covariate data, which can compromise model stability. Misclassification of CODs could arise from, e.g., recording errors, differing case definitions and causal hierarchies across studies, or poor interviewee recall in VAs; covariate measurement error can arise from imprecise measurements of difficult-to-measure metrics.

## 2. Proposed new methods

Various statistical methods are available which can address each of the above limitations, but implementing them within the existing classical (frequentist) framework proved challenging.

A mixed effects (ME) model with random country-specific intercepts is a way to give more weight to country-specific empirical data. These models are based on a hierarchical structure that assumes that some parameters do not vary (i.e. the fixed effects [FE] component) while others are treated as random variables (i.e. the random effects [RE] component) (Snijders, 2005).

Further, regularization techniques are a promising set of methods to simultaneously address the covariate selection and stability issues, by placing a penalty on model complexity. Most of these methods focus on two ways in which instability arises in the context of out-of-sample predictions: 1) increasing the number of covariates increases model complexity and therefore the risk of overfitting the model to the data; and 2) large coefficient values can increase instability. A subset of regularization methods exist that enable covariate selection within a multinomial framework without being computationally prohibitive. Least Absolute Shrinkage and Selection Operator (LASSO) and ridge regressions are examples of such regularization methods (James et al., 2013). Their general approach is based on including all covariates in the model and maximizing the log-likelihood minus a penalization/regularization term, which is a function of the model coefficients. This results in coefficient values with reduced magnitude. Increasing the penalization term in the LASSO regression pushes some covariates to zero (or very close to zero), hence performing a type of covariate selection within the multinomial model itself. We attempted to implement the LASSO regression within our frequentist multinomial modelling framework by adding a penalty term to the likelihood function. The implementation appeared to work in terms of shrinking covariate coefficients towards zero as the LASSO penalty increased in value. However, we consistently ran into convergence problems.

To address these implementation challenges, we propose shifting our multinomial logistic regression model from a frequentist framework implemented in Stata (www.stata.com) to a Bayesian framework in R (R Core Team, 2020), incorporating both country-specific random effects and the Bayesian LASSO for covariate selection.

### 2.1. Shifting to a Bayesian framework

The Bayesian framework is well suited to address the challenges discussed above. First, while mixed-effect models for multinomial logistic regression have been developed for the classical framework, their flexibility is limited (Hedeker, 2003). A similar model for multinomial data has been developed from the Bayesian perspective (Albert and Chib, 1993) and has been widely used (Burda et al., 2008; Jostins and McVean, 2016) and adapted for specific scenarios such as data sparsity (Cawley et al., 2007) and high dimensions (Yau et al., 2003). Moreover, adding in random effects and implementing the LASSO are both straightforward in the Bayesian framework through the use of priors and Markov chain Monte Carlo (MCMC) sampling from the posterior distributions.

We addressed two key methodological issues in order to implement the Bayesian neonatal COD models. First, we update our method of dealing with unreported CODs (Sect. 1.2) by specifying a matrix that can be incorporated as data in the modelling framework, so that (as before) all studies can be included in the multinomial model. Second, we implement the Bayesian LASSO with selection of the penalty term *λ*. Approaches exist to select *λ* during model estimation (Park and Casella, 2008), but since our model is designed to make out-of-sample predictions we require an alternative approach that maximises out-of-sample GOF.

### 2.2. Derivation of Bayesian multinomial model with random effects

Our proposed statistical model has several components including the basic multinomial model, the misclassification matrix, and the LASSO penalization term. These are described below.

#### 2.2.1. Basic model

Suppose there exist *C* mutually exclusive causes of death, and that we have a sample of *N*_*s*_ deaths from a given study *s*, each of which is (correctly) classified into one and only one of the *C* categories. If we denote the distribution of true CODs in the sample (i.e. our eight key causes) as *T*_1,*s*_, *T*_2,*s*_, …, *T*_*C,s*_ and if the sample is random, we can assume that these observations come from a multinomial distribution,

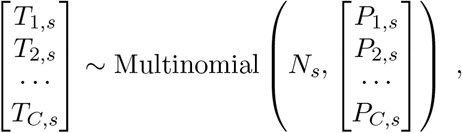

where *P*_*c,s*_ represents the probability that a death is due to cause *c* in the population in which study *s* is conducted. This can be re-written as

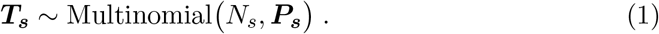

Because the *C* causes are mutually exclusive, it follows that 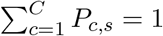.

#### 2.2.2. Misclassification matrix

Non-reporting of CODs can occur in studies as described previously and, to make matters worse, patterns of non-reporting may differ across studies. However, we can deal with this by specifying unreported causes as parts of residual causes. For each study, there is a specific misclassification matrix with the general form

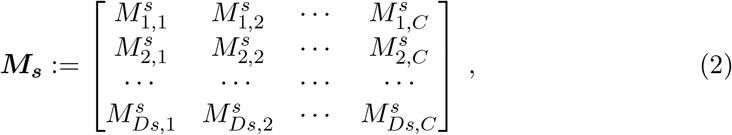

where 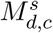 is the probability that, in study *s*, a death from *true* cause *c* is *recorded* as being due to cause *d*. Since studies record only one cause per death, each column of this matrix must add up to 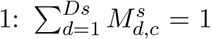 for all *c* = 1, …, *C*. The number of different *recorded* causes of death in this study is *Ds* and this can vary between studies. In fact, studies might also differ in the type of recorded causes of death, not only in the number of these. Some recorded causes might appear only in some studies.

Using (2), for a given study *s* we can express the recorded COD multinomial probability distribution as ***M***_***s***_ *×* ***P***_***s***_, where ***M***_***s***_ is the study-specific misclassification matrix and ***P***_***s***_ is the study’s true probability distribution. Because ***M***_***s***_ is not necessarily invertible, we cannot directly estimate ***P***_***s***_ from the probability distribution of recorded causes. Nevertheless, we can still use the distribution of the recorded causes of death along with the misclassification matrix to estimate the model coefficients, as follows.

#### 2.2.3. Proposed model

Suppose the probabilities *P*_*c,s*_ can be predicted by the values of a set of *K* explanatory variables *X*_1,*s*_, *X*_2,*s*_, …, *X*_*K,s*_. In a multinomial regression framework we assume that the logarithm of the odds of each cause of death relative to a reference cause are linearly dependent on these explanatory variables. This is expressed as a system of *C −* 1 linear equations corresponding to each cause of death (excluding the reference category),

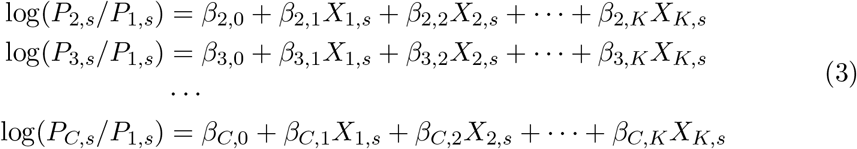

Notice that the *β*-coefficients (including the intercepts) do not have the study subindex *s*. This is a fixed-effects model that assumes the associations of the explanatory variables with the causes of death are constant across all studies. We relax the assumption that the baseline log-odds are the same in each study by adding study-specific random effects to the intercepts. Using matrix notation, and including random effects, the model in (3) can be expressed as

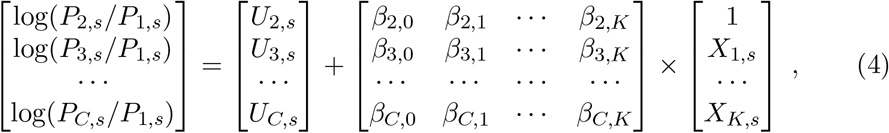

where the terms *U*_*c,s*_ are study-specific and can be modelled as random effects with mean 0 across all studies. The notation in (4) could be simplified to

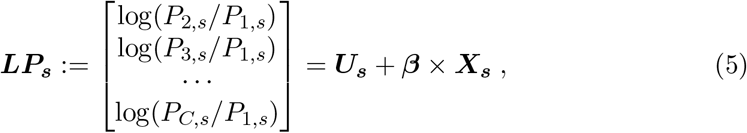

where ***U***_***s***_ and ***X***_***s***_ are the study-specific vectors of random effects and explanatory variables respectively, and ***β*** is the matrix of fixed effects common to all studies.

### 2.3. Bayesian LASSO

In a Bayesian framework, we implement LASSO covariate selection by penalising large *β* coefficients in a subset of the fixed-effect parameters that could potentially result in overfitting the data. We do this by imposing a double exponential (also referred to as Laplace) prior distribution on them in the model specification (Park and Casella, 2008). Unlike the frequentist LASSO, the Bayesian LASSO shrinks the magnitude of the parameters without completely reducing them to zero, allowing covariates to have negligible effects for some outcomes and non-negligible effects for others. Shrinking the parameters has the additional advantage of stabilising the model if, due to the large number of parameters to be estimated with potentially high uncertainty, model convergence is slow or difficult.

#### 2.3.1. Formal model definition

Our proposed method specifies Eq. (5) in the statistical model and uses an MCMC sampling algorithm to build it in a Bayesian framework. Note that the vector of true causes of death is unobserved, but with a specification for the true cause distribution, a vector of observed reported causes and a (known) misclassification matrix, we can specify a multinomial distribution for the observed reported causes. Let *N*_*s*_ denote the sample of deaths from a given study *s*, and ***M***_***s***_ the misclassification matrix defined in (2), our Bayesian model can be summarised as follows:

LIKELIHOOD:

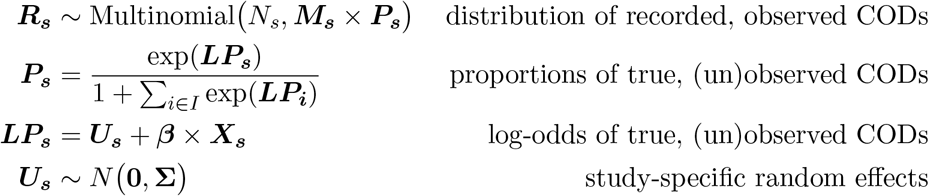

PRIORS:

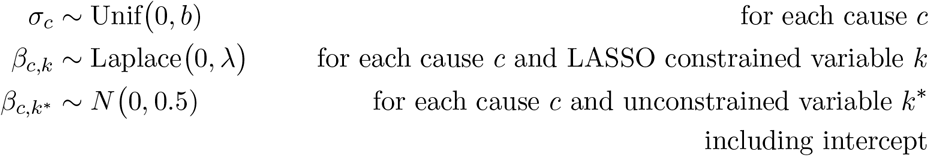

Note that ***R***_***s***_, *N*_*s*_ and ***X***_***s***_ are observed data, whereas ***M***_***s***_ is assumed to be known. Vector parameter **Σ** = (*σ*_2_, …, *σ*_*C*_) contains *C −* 1 standard deviations of the random effects, one for each cause except for the reference category. These standard deviations have uniformly distributed priors controlled by hyper-parameter *b*. For simplicity, we assumed there is no correlation between random effects of different causes, although this could be modelled in other ways. The intercepts and any *β*-coefficients we do not want to be constrained in the LASSO are given normally distributed priors with mean 0 and standard deviation 0.5. The remaining *β*-coefficients have a Laplace (double exponential) prior with mean 0 and precision *λ >* 0. The hyper-parameter *λ* is the penalty imposed by the LASSO method, giving a standard deviation of 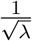. We use out-of-sample cross-validation to select optimal *λ* and *b* parameters, described in the next section.

#### 2.3.2. Estimation of country-level COD mortality fractions with credible intervals

Once the model has estimated ***β*** and ***U***_***s***_ correcting for potential misclassification we can estimate the expected distribution of true COD in any country for which we have covariate data as

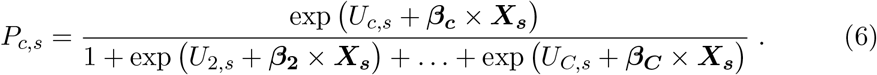

using the posterior means of the fixed and random-effects coefficients.

When using this model to estimate country-level COD fractions we need to account for two sources of uncertainty: a) uncertainty surrounding the fixed-effects parameter estimates; and b) uncertainty about how to select the most appropriate random effect *U*_*c,s*_ for the country that we want to estimate, particularly if that country was not represented in our input data. In a Bayesian framework, credible intervals around the COD fractions can account for both sources of uncertainty. We compute these credible intervals by repeatedly estimating the COD distribution using values of *β* and relevant (described below) random effects drawn from MCMC chains generated during model estimation. We use *n* = 1,000 sets of estimates from equally spaced MCMC iterations after burn-in, although the selected sets can also be determined by a thinning parameter or even randomly selected. We thus obtain *n* sets of estimated mortality fractions for each country, from which we find the 2.5^th^ and 97.5^th^ centiles (to obtain 95% credible intervals) and mean and median values (to obtain point estimates) for each COD.

#### 2.3.3. Choice of random effects

An important question is how to select relevant random effects from the matrices ***U***^***i***^ in each iteration *i* = 1 … *n*, because the choice affects both the point estimates and the credible intervals. Each ***U***^***i***^ contains a vector of random effects for each study in the estimation dataset. There are several ways of deciding which is the most appropriate vector for a given country; we illustrate three below:

a. We could assume ***U***_***s***_ = 0 in all iterations, which may seem a sensible strategy for countries not represented in the input data. However, this is an extreme option representing a strong prior belief that the country’s COD distribution is exactly predicted by the model’s fixed effects, when no evidence suggests this. As such it will produce overly narrow credible intervals, with variability determined only by fixed effect estimate uncertainty. We do not explore this option further.
b. Choose a vector ***U***_***s***_ at random from the matrix in each iteration. This strategy relaxes the belief that the country’s COD distribution is exactly predicted by the model’s fixed effects by drawing from a wide range of random effects at each iteration. In expectation these random effects sum to zero, having little effect on the point estimate, but because many different random effects are drawn the uncertainty in the COD distribution estimate is large. We expect wide credible intervals with variability dependent on both fixed and random effect estimate uncertainty.
c. Choose a vector ***U***_***s***_ at random from a relevant subset of the matrix in each iteration. This is an intermediate option that assumes random effects from a certain subset of studies, which may not necessarily sum to zero, provide additional relevant information for the country’s predictions. This might move the point estimate away from the fixed effects predictions and will produce credible intervals of a width somewhere in between options (a) and (b), with variability determined as in (b) but incorporating less uncertainty in the selection of relevant random effects.

We implement options (b) and (c) to avoid understating the uncertainty in country-level predictions, particularly for countries in which we have no nationally representative data. Thus, for countries with no studies or only non-nationally representative studies in the input data, we use option (b) to obtain wide credible intervals with little effect on point estimates. For countries with nationally representative studies, we use option (c) drawing from a subset of ***U***^***i***^ containing only their nationally representative random effects. This gives narrower credible intervals than option (b), acknowledging that we have more confidence in these estimates, and may move the point estimates away from the fixed-effects means.

## 3. Model building process

### 3.1. Misclassification matrix

We specified the misclassification matrix ***M***_***s***_ separately for each study using the logic from our previous method for handling unreported CODs (Sect. 1.2). We recorded deaths that were reported as one of our eight key causes with 100% probability as that cause by placing a 1 in the relevant cell of ***M***_***s***_. Unreported deaths from intrapartum, preterm, congenital and sepsis causes were assumed to have been recorded under ‘other’ causes of death, and we placed an additional 1 in the ‘other’ row of ***M***_***s***_ in the cell corresponding to the relevant column for the missing cause, thus representing a 100% probability that these deaths were recorded as ‘other’. Unreported deaths from pneumonia, diarrhoea and tetanus were first assumed to have been reported as ‘sepsis’, if ‘sepsis’ was reported in the study, or as ‘other’ if not, and we placed 1s in the ‘sepsis’ or ‘other’ row, respectively, following the same procedure. Using this method, each column, representing our key CODs, contained a single 1 and each row, representing studies’ reported CODs, contained one or more 1s. The maximum number of unreported causes was four, from one study, where pneumonia, diarrhoea, tetanus and sepsis deaths were unspecified, and we assumed they were reported under ‘other’.

### 3.2. Model building process

We used the same 95 studies from the previous round of frequentist modelling of neonatal COD estimation (Sect. 1.1) (Oza, 2019) in fitting the Bayesian model. Sixteen of the studies were nationally representative of the country in which they were conducted (**Figure e1, Online Supplement**), and 10 came from countries that were still considered high mortality during the prediction period (2000-2015, see Section 4.3). As before, we let “intrapartum” be the reference COD in the multinomial model and build equations for the other seven causes (detailed in Sect. 1.2) compared to “intrapartum”. Each model equation contained an intercept term, 14 covariates (Sect. 1.1) and a random effect term, all of which were allowed to vary by COD.

#### 3.2.1. Fixed effects

We modelled all covariates linearly, with no polynomial effects or interactions. The beta matrix therefore contained 105 coefficients for estimation.

The early and late period indicators (as well as the intercept) for each COD (21 coefficients) were given *N* (0, 0.5) priors free from the LASSO penalisation to ensure the two mortality periods can have different COD distributions where appropriate. The remaining 84 coefficients were each subject to LASSO by imposing the Laplace(0, *λ*) prior. We explored *λ* values of 5, and 10 to 250 in increments of 20. This wide range allows for different values of *b* to suggest different optimal values of *λ*.

#### 3.2.2. Random effects

We tagged each study with an indicator variable for the random effects term and identified whether it was nationally representative. This resulted in 95 sets of random effects, and the U matrix therefore contained 665 random effect coefficients for estimation.

We gave the random effects distributions *N* (0, *σ*_*c*_) priors, and gave each *σ*_*c*_ the hyper-prior Unif(0, *b*). The hyper-prior controlled by *b* imposes a ceiling on the random effect standard deviations that helps the model to avoid overfitting random effects to noisy data, which could potentially eliminate the predictive value of fixed effects, and limits the extent to which study data can influence country predictions. We impose a hard upper limit on *b* to avoid drastic differences in mortality fractions between two countries with the same epidemiological situation (same levels of predictive covariates), but which provided different study-level evidence. For example, when *b* = 0.21 only 5% of random effects in a particular cause’s posterior distribution should affect the fixed-effects odds for that cause by a magnitude greater than 1.51 or its reciprocal. We let this be the upper limit of influence and explored two more restrictive constraints of *b* = 0.07 and *b* = 0.14.

### 3.3. Formal selection of λ and b using cross-validation

We selected *λ* and *b* jointly by partitioning the dataset into *k* subsets and performing a *k*-fold cross-validation over a selection of values of both parameters. With *k* = 10 we found that the error curves were sensitive enough to the choice of the partition to suggest different values for *λ*, providing evidence for high variability but (because *k* is low relative to sample size) no theoretical guarantee of unbiasedness. Instead, we therefore used leave-one-out cross-validation, in which the error curve is theoretically highly variable but approximately unbiased (Jiang et al., 2002). We set *k* at 95, the number of studies in our dataset.

For each combination of *λ* and *b*, we ran a full leave-one-out analysis in the following way: we remove a study from the input data, run the MCMC model with the remaining 94 studies and predict the distribution of deaths in the out-of-sample study using the posterior means of the fixed effects. We ignored random effects at this stage because, using the scheme we describe in Section 2.3.3, there were no suitable random effects to use in a single out-of-sample estimate of the COD distribution.

We repeat this 95 times, leaving a different study out each time so that we obtain an out-of-sample predicted COD distribution for each left-out study. For each study and COD we calculate the squared differences between predicted and observed deaths, weight them by the ratio of the predicted deaths to the total deaths over all studies and CODs, and sum these errors over all studies and CODs to obtain a weighted mean square error (wMSE) for each jackknife sample. We compare the wMSE across jackknife samples (over different values of *λ* and *b*) and select the combination that produces the lowest absolute value.

#### 3.3.1. Parameters for the MCMC convergence

To avoid the effects of autocorrelation and ensure convergence, we ran models in four parallel chains of 10,000 iterations each and an initial burn-in sequence of 2,000. We used trace and Gelman plots (Brooks and Gelman, 1998) of the coefficients to determine the number of iterations to burn, choosing the iteration at which the slowest parameter had converged over all four chains (flat locally weighted regression lines on MCMC trace) and the chains showed evidence of convergence (scale reduction factor below 1.1 over majority of iterations). We determined the number of post-burn iterations from the parameter with the strongest autocorrelation, ensuring that the four trace lines cycled through the posterior median at least twice beyond the burn point. For this reason we did not thin our posterior distributions: biasing effects of autocorrelation should average out, given the large number of iterations and chains.

Further modelling details are available in **Online Supplement Text E2**. We used R v3.5.2 for all analyses (R Core Team, 2020). Bayesian analyses were implemented in JAGS (Plummer, 2003) with wrapper functions from the R2jags package (Su and Yajima, 2020).

Statistical code is available on https://github.com/amulick/MCEE-neo.git.

## 4. Results and evaluation

We ran leave-one-out models on a 2.8 gigaHertz (GHz) machine with eight parallel processors, each of which completed in ∼ 16.5 hours using approximately 26 GB of RAM. Total computation time including final model estimation was approximately 700 hours. Further details are available in **Online Supplement Text E3**.

### 4.1. Cross-validation

We ran leave-one-out analyses at each of the 42 combinations of *λ* and *b*. Figure 1 plots the out-of-sample wMSE against values of *λ* over three values of *b*. The initial decline of these curves is a reflection of the overfitting of the fixed effect coefficients of the model (i.e. the effect of the covariates used for prediction) to the input data, and the later incline reflects underfitting; thus the value of *λ* at the minimum wMSE indicates the model with the best out-of-sample fit. These curves were steeper and had lower minima at higher *b* values, such that the most accurate predictions were achieved with the least restrictive cap on *σ*_*c*_ (*b* = 0.21).

**Fig. 1.**
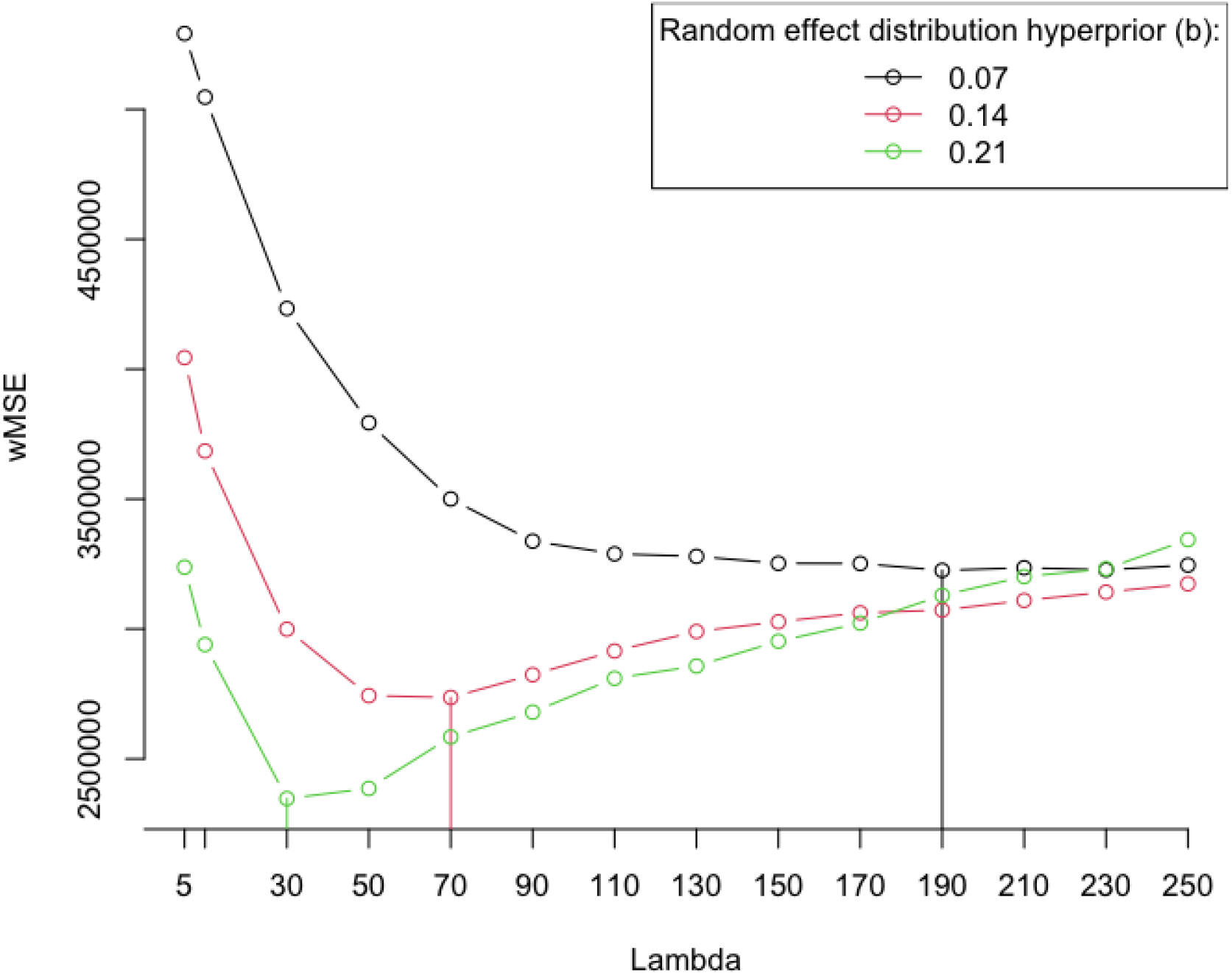
Weighted Mean Square Error (wMSE) from out-of-sample neonatal cause-of-death predictions plotted against values of fixed-effects shrinkage parameter *λ* over three values of random effect distribution hyper-priors *b*

This illustrates that the selection of the best predictive model depends on both *λ* and *b*. Among the combinations of *λ* and *b* examined, we obtained the worst out-of-sample prediction (wMSE *>* 5 × 10^6^) when the fixed effects are most flexible by both parameters (*λ* = 5, *b* = 0.07) – a clear example of overfitting. As we restrict the magnitude of the fixed effects by increasing *λ*, the out-of-sample prediction initially improves over all three values of *b*. At the same time, as we “relieve” the fixed effects from having to fit the data by allowing the model to have larger random effects (increasing *b*; moving from the black to the red to the green line in Fig. 1), we also improve out-of-sample prediction. The combination of these two measures prevents overfitting, but eventually with large enough *λ* we restrict the fixed effect coefficients too much and predictions worsen again. This limit appears at lower *λ* if the random effects are allowed to explain more data variability with larger *b*.

### 4.2. Final models

To understand the differences between these three best-fit models, we ran a final model for each of them using all data points. Trace and Gelman plots for the covariates and nationally representative random country effects showed good convergence in all three models. Most of the seven *σ*_*c*_ parameters were sampled consistently at or near *b*.

The difference in LASSO-constrained beta estimates was generally small (Fig. 2), although some estimates were notably weaker in the *λ* = 190 and *b* = 0.07 model compared to the other two models. In this model “other” causes of death, compared with other CODs, show greater differences particularly in the GFR and SBA coefficients. These differences balance in the unconstrained coefficients: e.g., the GFR and SBA coefficients are estimated less strongly positive than the other two models, but the unconstrained coefficient “per.early” is estimated more strongly negative than the other two models, which is more easily visible when viewed on the same scale. This highlights the interaction between *λ* and *b* described in Sect. 4.1.

**Fig. 2.**
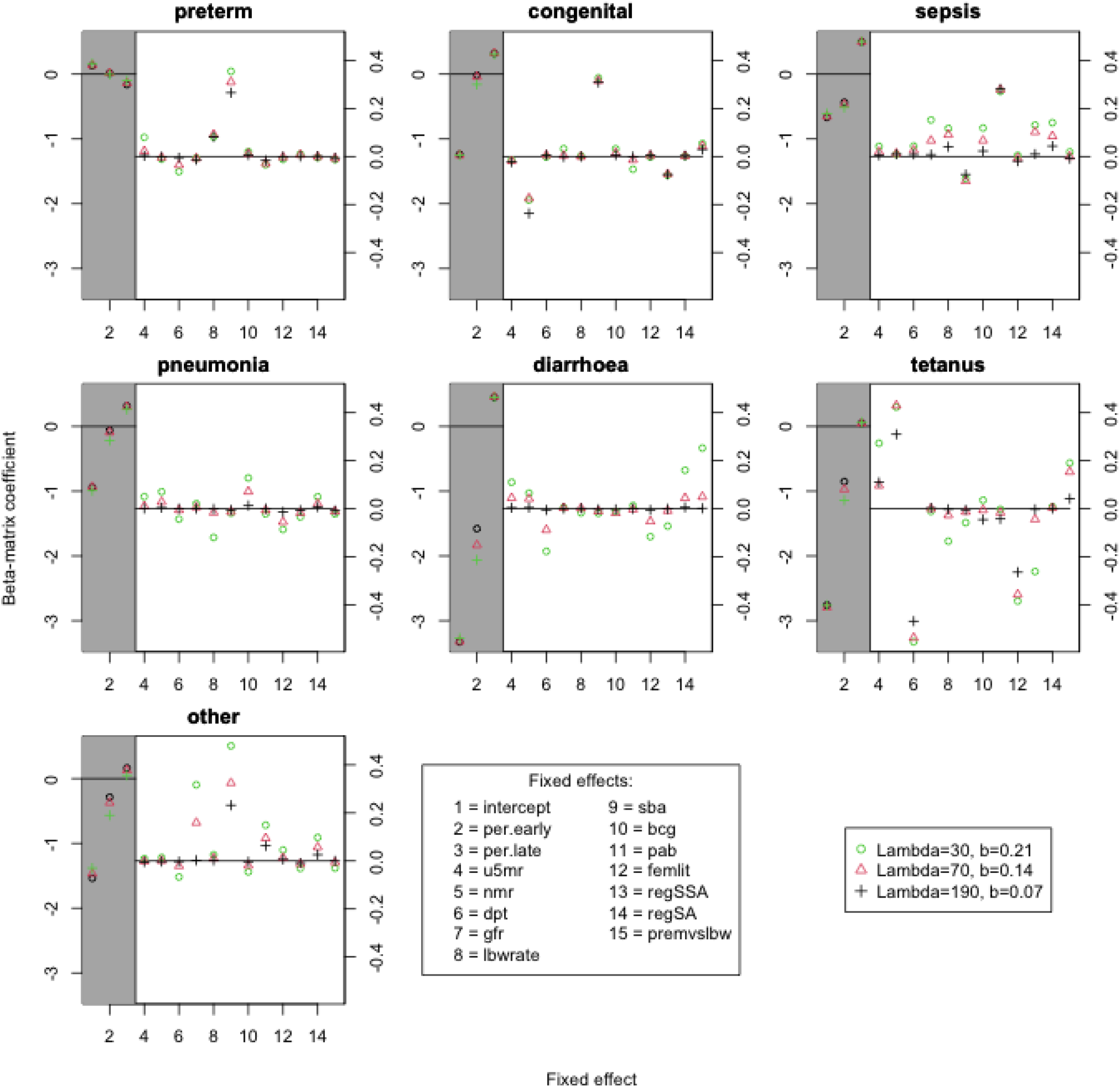
Bayesian fixed effects coefficients compared between three neonatal cause-of-death models with different values of fixed-effect shrinkage parameters *λ* and random effect distribution hyper-priors *b*

The difference in random effects was greater (Fig. 3) among the three models. Although some estimates were similar between them, most were markedly different with, as expected, larger effects appearing in models with larger values of *b*.

**Fig. 3.**
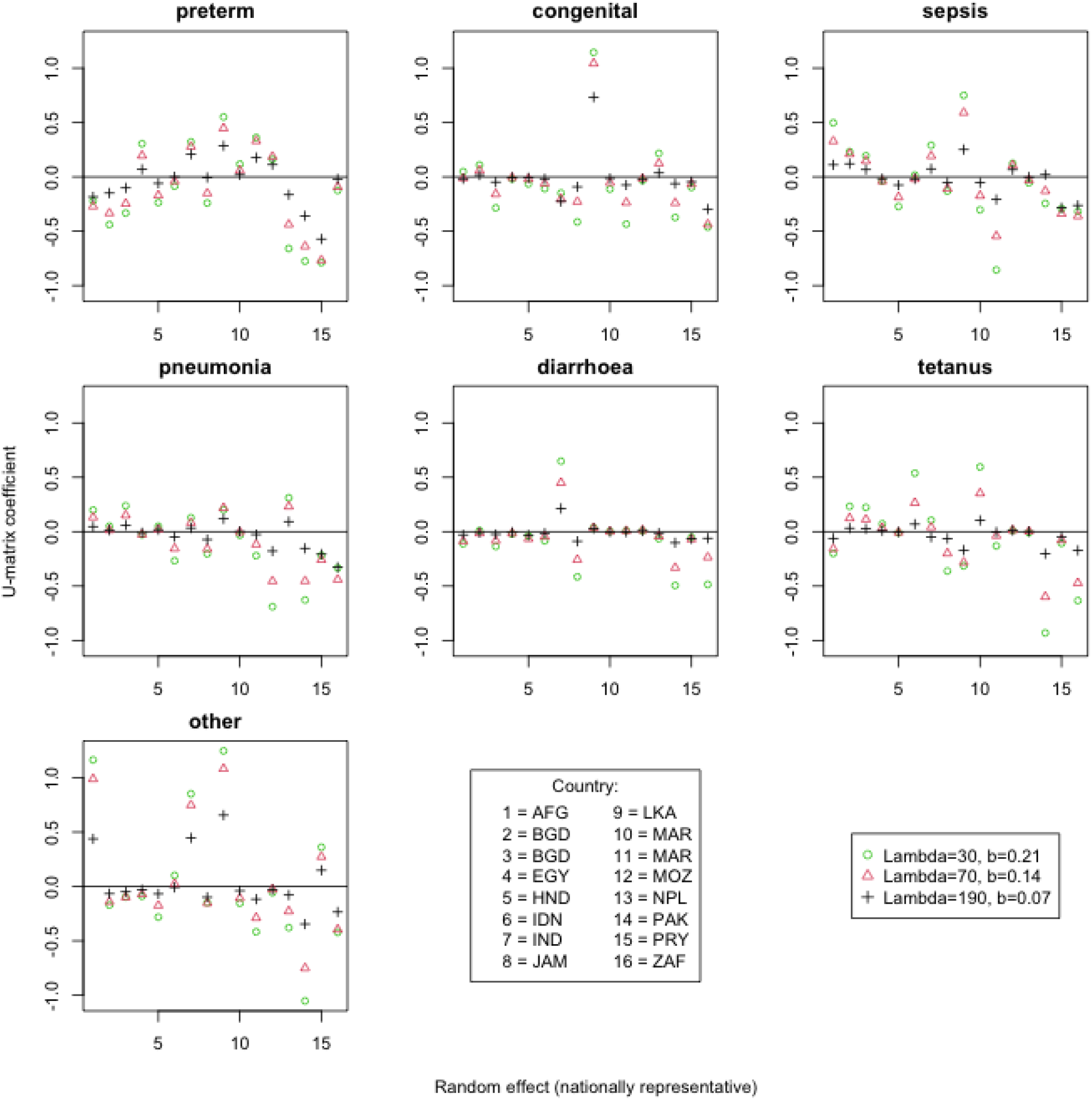
Bayesian random effects coefficients compared between three neonatal cause-of-death models with different values of fixed-effect shrinkage parameters *λ* and random effect distribution hyper-priors *b*. Sixteen sets of random effects are from the studies with nationally representative input data for their country

### 4.3. Predictions

We used the model with the best out-of-sample fit (*λ* = 30 and *b* = 0.21) to predict COD distributions in 80 countries between the years 2000 and 2015 inclusive. For 8 of the 80 countries nationally representative data were available: Bangladesh and Morocco had two nationally representative studies and Afghanistan, India, Indonesia, Mozambique, Nepal and Pakistan had a single study. Unless otherwise noted, to make comparisons easier we present results in this section using single years rather than the full 16-year prediction period.

#### 4.3.1. Frequentist vs Bayesian

Figure 4 and Table 1 compare predicted proportions of all causes of death between the classical frequentist model and our proposed Bayesian model (fixed effects only) in 2015. This provides comparable estimates in that the proportions differ only in the method used to develop the predictive model.

**Fig. 4.**
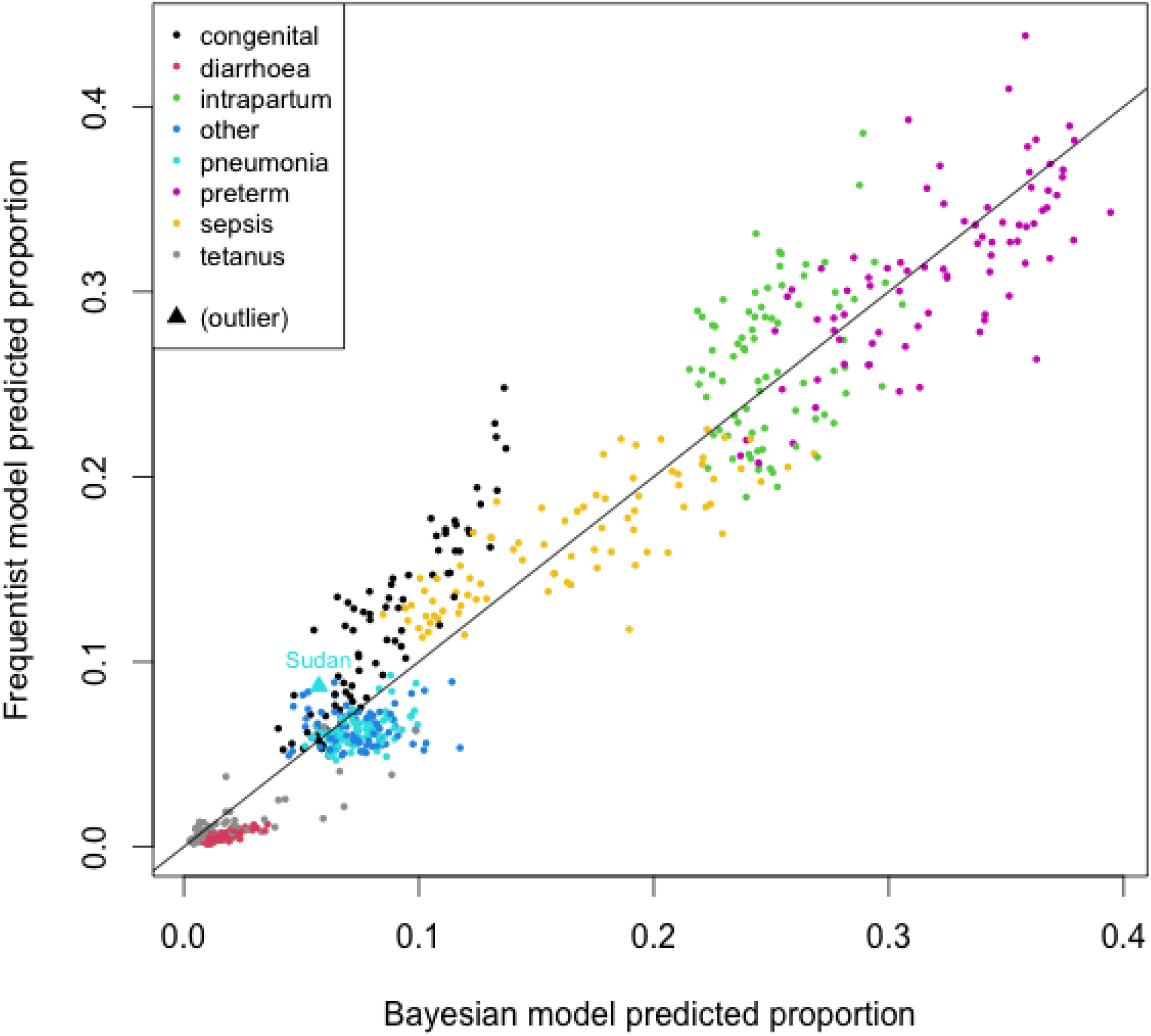
Predicted Bayesian fixed-effects cause-of-death proportions (posterior means) compared with predicted frequentist proportions in 2015 for 80 countries

**Table 1.** Differences in predicted cause-of-death fractions for 2015, Bayesian fixed-effects posterior means compared to frequentist fixed-effects estimates, for 80 countries

| COD | Absolute difference: Median [IQR] | Relative difference: Median [IQR] |
| --- | --- | --- |
| congenital | 0.03 [−0.05, −0.01] | 0.73 [0.64, 0.82] |
| diarrhoea | 0.01 [0.01, 0.01] | 3.24 [2.80, 3.68] |
| intrapartum | −0.02 [−0.04, 0.02] | 0.94 [0.85, 1.03] |
| pneumonia | 0.01 [0.00, 0.02] | 1.17 [1.03, 1.31] |
| preterm | 0.01 [−0.01, 0.03] | 1.04 [0.97, 1.11] |
| sepsis | −0.01 [−0.02, 0.02] | 0.93 [0.85, 1.01] |
| tetanus | 0.00 [0.00, 0.01] | 1.59 [1.05, 2.13] |
| other | 0.01 [0.00, 0.02] | 1.16 [1.02, 1.30] |

Predicted proportions for intrapartum, preterm and sepsis were distributed roughly equally on either side of the line of equivalence; for the other causes one of the models tended to predict larger proportions than the other. The median difference in absolute terms was largest for congenital proportions (Bayesian estimates 3% lower than frequentist); in relative terms the difference was greatest for diarrhoea (Bayesian estimates on average 3 times higher). However, both models assigned relatively small proportions (maximum 0.25) to both of these causes.

Within each cause we highlight outliers (countries whose estimate is greater than 3 standard deviations from the average difference between the two methods) with a triangle. There was only one such difference: Sudan’s Bayesian pneumonia estimate was markedly lower than its frequentist estimate.

#### 4.3.2. Bayesian: Fixed vs random effects

Figure 5 compares point estimates from fixed-effects only and fixed-plus random-effects predictions for countries with nationally representative input data (except Morocco, because its studies preceded 2000). For most countries and CODs, the fixed-plus random-effects predicted proportion lies between the empirical proportion than the predicted proportion based on the fixed effects only. There were some exceptions: 1) Bangladesh (2002), congenital; 2) India, diarrhoea; 3) Mozambique, intrapartum and other; 4) Nepal, diarrhoea and tetanus; and 5) Pakistan, congenital. For these, the estimates incorporating the random effects were all further away from the empirical proportion than the estimates based on the fixed effects only.

**Fig. 5.**
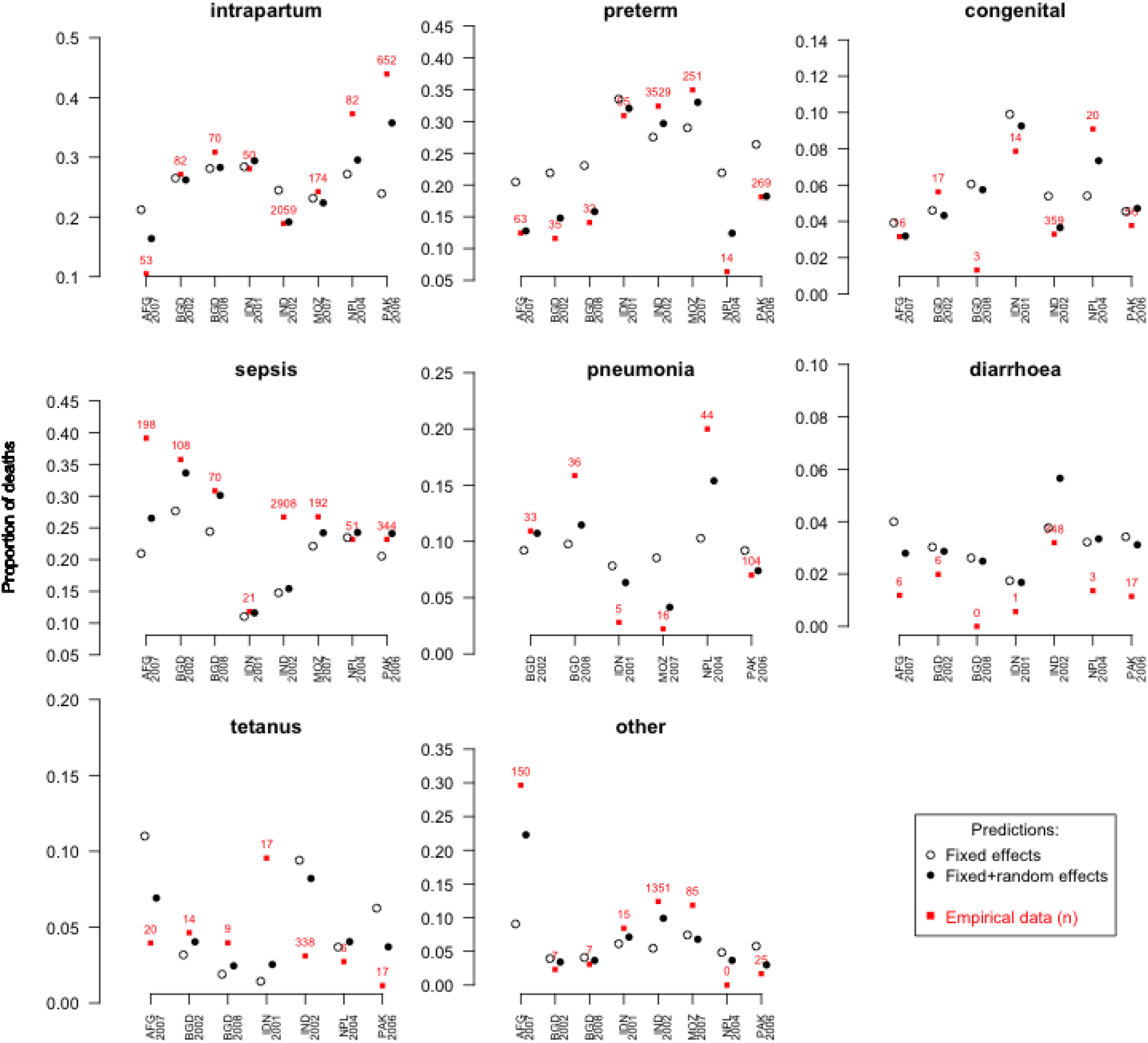
Fixed-effects only versus fixed-plus random-effects estimates of cause-of-death distributions in countries contributing nationally representative data to the Bayesian model. Red squares mark the study COD proportion and indicate the absolute number of deaths it represents. Bangladesh is represented twice because it provided two nationally representative studies. Afghanistan, India and Mozambique do not appear in all causes because their studies did not report deaths in the same categories that we report

The first exception occurred because Bangladesh has two empirical data points (2002 and 2008), both of which informed its random effects, and these fell on either side of the fixed-effects prediction for congenital. Bangladesh’s fixed-plus random-effects prediction equation drew randomly from both random effects to produce the posterior distribution of point estimates, so that the mean prediction averaged out the two random effects. For all other CODs, Bangladesh’s empirical data points were on the same side of the fixed-effects estimates.

The second and third exceptions occurred because of non-reporting of key CODs. India did not report pneumonia deaths and Mozambique did not report congenital, diarrhoea or tetanus deaths, so the misclassification matrix for these studies redistributed some of the deaths they did report into these causes. In this way, the random effects for each model COD are not expected to necessarily pull all proportions in the same direction as the empirical data.

The fourth and fifth exceptions likely occurred due to chance. The average random effects (Nepal: diarrhoea and tetanus, Pakistan: congenital) were all very close to zero and the change in predictions due to adding the random effect were very small (Fig. 5). Thus the slight pull in the opposite direction was likely due to the prediction algorithm sampling, by chance, more random effects on the opposite side of zero than the empirical data would suggest.

In general, though not exclusively, the empirical proportions representing larger numbers of deaths pull the random-effects estimates more strongly towards themselves than proportions representing fewer deaths. This can be seen by comparing Nepal (2004) and Pakistan (2006): Pakistan’s study reported on larger numbers of deaths and their random-effects predictions for nearly all CODs are closer to their empirical data than Nepal’s predictions are to their empirical data.

### 4.4. Uncertainty ranges

Figure 6 compares the 2015 mortality proportions and uncertainty ranges across four causes of death between the classical frequentist model and our proposed Bayesian model for countries with the highest burden of neonatal mortality and/or nationally representative input data. This shows the joint effect of moving to a Bayesian framework and adding random effects into the model in countries where the differences in number of deaths would be most extreme. Most estimates from countries without nationally representative data are close to the frequentist point estimates, or within the uncertainty limits of the frequentist estimates. In general, the Bayesian credible intervals are more symmetric around the point estimates than the bootstrapped frequentist confidence intervals.

**Fig. 6.**
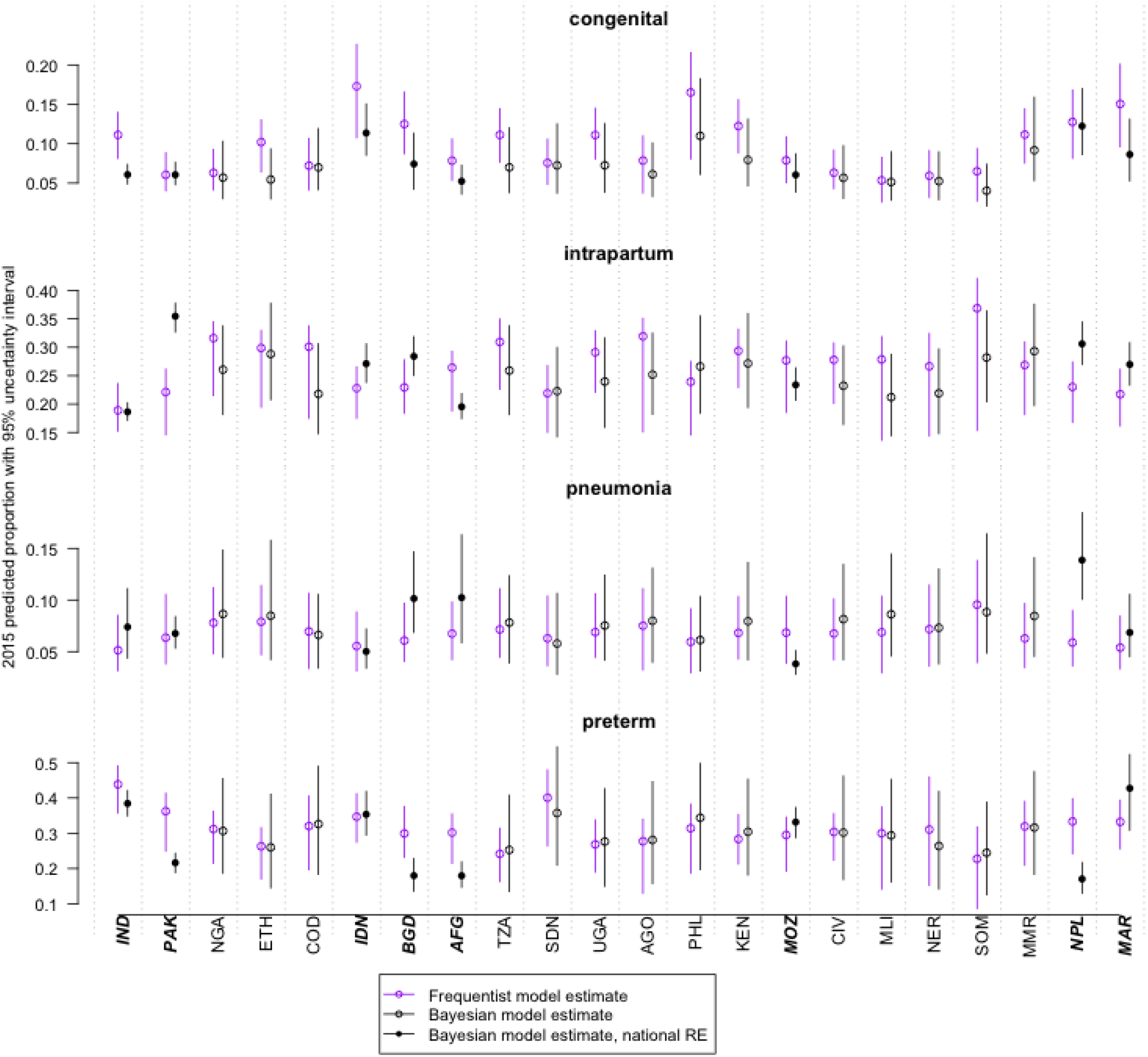
Mortality proportions and uncertainty ranges for select neonatal causes of death in 2015 compared between a classical frequentist model with bootstrapped confidence intervals and our proposed Bayesian model (fixed-plus random-effects estimates) with credible intervals. In descending order from left to right are a union of 20 countries with the highest burden of neonatal mortality in 2015, and eight countries that contributed nationally representative studies to the Bayesian model. Countries contributing nationally representative data are highlighted in bold italic

Table 2 summarises, for 80 countries with available data, differences in the widths of the uncertainty intervals. For countries without nationally representative data, the Bayesian intervals were generally wider than the frequentist, particularly for pneumonia and preterm CODs.

**Table 2.** Differences in uncertainty between our proposed Bayesian model, including fixed and random effects to make the predictions, and the classical frequentist model (fixed effects only) for selected 2015 COD fractions in 80 countries. The widths of bootstrapped frequentist confidence intervals are subtracted from the widths of Bayesian credible intervals. Values greater than zero indicate how much wider the Bayesian intervals are, in absolute value, than the frequentist intervals

| COD | *n* | Median [IQR] |
| --- | --- | --- |
| Countries without nationally representative studies |  |  |
| congenital | 72 | 0.01 [0.00, 0.02] |
| intrapartum | 72 | 0.02 [−0.01, 0.05] |
| pneumonia | 72 | 0.02 [0.01, 0.03] |
| preterm | 72 | 0.11 [0.08, 0.14] |
| Countries with nationally representative studies |  |  |
| congenital | 8 | −0.02 [−0.03, −0.01] |
| intrapartum | 8 | −0.04 [−0.06, −0.02] |
| pneumonia | 8 | 0.01 [−0.02, 0.04] |
| preterm | 8 | −0.06 [−0.07, −0.05] |

In contrast, estimates for the countries that did provide nationally representative data generally have narrower Bayesian than frequentist uncertainty intervals. This is because the algorithm calculating the fixed-plus random-effects estimate is drawing from only one or two relevant random effects, rather than all 95 random effects. Fewer random effects add less variability to the posterior distribution; thus its 2.5^th^ and 97.5^th^ centiles are closer together. This has the initially counterintuitive consequence that countries providing more nationally representative data, such as Bangladesh and Morocco with two studies, can have wider uncertainty intervals than they would had they provided only one study.

## Discussion

We developed a new approach within a Bayesian framework for estimating the distribution of causes of death at national level based on COD data from national and subnational studies and data on covariates. Compared with the previous frequentist approach, this approach resulted in similar estimates on average for the most common causes of death, but some differences in the estimates for the less common causes. The incorporation of random effects into the model led to somewhat increased levels of uncertainty, with the exception of countries with nationally representative studies, where estimated uncertainty was not unexpectedly decreased. We believe that the wider uncertainty ranges generated under the new approach better reflect real uncertainty that exists given the substantial scope for misclassification in verbal autopsy studies.

By working within the Bayesian framework we were able to incorporate features which we could not in a frequentist setting. First, we were able to better incorporate empirical data from nationally representative studies through the use of random effects, so that estimates for countries with empirical data from nationally representative studies are “pulled towards” those empirical data. Second, we were able to examine the associations between a set of predictors and all causes of interest simultaneously with the Bayesian LASSO, rather than selecting predictors in a more na ï ve process with each cause individually or in an exhaustive and computationally prohibitive search across predictors in the multinomial space. Third, we were able to recast our method for mapping between causes as they are reported and specific causes of interest (the misclassification matrix), opening up possibilities for future extensions discussed below.

Including random effects for countries with nationally representative studies has several important advantages. Countries with nationally representative cause-of-death information contribute to estimates for all countries through the model’s fixed effects coefficients, but also have greater influence on the estimates for that country which likely reduces bias (Bouwmeester et al., 2013; McCulloch and Neuhaus, 2011). This also is intuitively a pleasing compromise between what countries report, which may have limitations (Menéndez et al., 2020), and what would be expected given the evidence from other areas with similar levels of mortality, intervention coverage, and other health system characteristics. To our knowledge there are not other methods that allow for such a systematic compromise in estimating causes of mortality. Allowing nationally representative cause-of-death measurement to have more influence may also encourage the collection of such data, which would increase the accuracy and usability of future estimates as well as the capacity in low-resource countries for such measurement.

We also incorporated cross-validation to determine the degree of penalization in the Bayesian LASSO. Although the Bayesian LASSO is often implemented with a fixed restriction or with a hyper-prior for the restriction parameter (Park and Casella, 2008), we chose the degree of restriction according to the cross validation error for more robust out of sample prediction (Efron and Tibshirani, 1995).

In addition to its advantages, the proposed method in the Bayesian framework is more flexible than the previous method in the frequentist setting, which we expect to allow for further improvement. For example, cause of death ascertainment can be prone to measurement error, particularly when using verbal autopsy (World Health Organization, 2016). With our new approach, this has potential for resolution through an extension to the misclassification matrix. This framework may also allow increased accounting for uncertainty and measurement error in the covariates for primary data, which often is not available at the same level of resolution for which the causes of mortality were measured (Liu et al., 2016).

We used credible intervals to estimate the uncertainty in predicted cause distributions for all countries by resampling from the MCMC-estimated random effects for each study, and among only those estimates from nationally representative studies for countries with such studies. This may in practice lead to the counterintuitive result that estimates from a country with more than one nationally representative study appear more uncertain than estimates from a country with only one nationally representative study. However, because studies from the same area may report different causes due to differences in study methods such as cause ascertainment (Murray et al., 2014) or due to epidemiologic changes over time, an increase in uncertainty may be warranted. Analogously, estimates from countries with a single nationally representative study may have uncertainty intervals that are too narrow, because they likely do not account for uncertainty due to possible error in the cause-of-death classification or variation in trends over time.

The proposed method is in contrast to those used by the Global Burden of Disease (GBD) consortium (Murray et al., 2020) for estimating the causes of mortality. This group looks at many separate causes for different age groups, including data from incomplete vital registration and registries for specific syndromes and aetiologies of disease. After all causes are estimated separately with Gaussian processes, they are then restricted to an age-specific envelope (total number of deaths due to all causes) in a separate process (Murray et al., 2020), although their methods and data sources are not publicly available in detail (Schwab, 2020). The method proposed here is not directly comparable to GBD methods because each are based on different source information. When approaching compositional data such as causes of death, addressing components individually as done by GBD is generally unbiased, but there are caveats for estimating the variance of separately estimated components which are subsequently fitted to an envelope (Begg and Gray, 1984; Fürnkranz, 2002; Hsu and Lin, 2002). The estimated variance for each component is used in the GBD method for harmonizing the many causes to fit the mortality envelope (Murray et al., 2020) and so may introduce bias in the resulting estimates. The vast amount of input data used by GBD allows for the estimation of many different causes, which would be computationally difficult in the multinomial framework. However, the “squeezing” process made necessary by the many different causes may lead to inaccuracies. The method proposed here is computationally complex, but it is executed in a single systematic framework that is used widely in other similar problems with compositional measures (Haan and Uhlendorff, 2006) and with attractive statistical properties that are well documented (Engel, 1988).

The proposed method is, as with many statistical methods, limited by the quality of primary measurements. Causes of death in high mortality and low resource settings are often subject to specific types of measurement error (Adewemimo et al., 2017). Both the amount of information related to causes of death in areas without vital registration (Datta et al., 2020) as well as the methods for measuring causes of death in such areas (Kalter et al., 2020; McCormick et al., 2016) are improving. However, historic cause-of-death measurements are likely unique, as health systems change and mortality among neonates and children declines. So, historic causes of mortality may need bespoke measures when attempting to correct for them (Yadav and Arokiasamy, 2014), which our extension to the misclassification matrix has potential to do. Another important limitation of the proposed method is the amount of time and computational resources necessary for implementation. We used parallel computing on a multi-node high performance computing cluster for these analyses. Such resources are not widely available and are associated with both financial and time related costs, as an analyst must learn both the method and the protocol of the computing cluster. High performance computing, however, is becoming more accessible and may be less of a barrier in the future (Clark, 2020).

Although there have been important advances in vital registration as well as sample registration systems for measuring causes of death for all individuals, there are still gaps in these systems that will likely make modelling causes of death necessary for the foreseeable future (Amouzou et al., 2020). Advances in the accuracy of cause-of-death estimates translate into better knowledge of what contributes most to age-specific mortality and how health systems can be configured for the biggest impact (Walker and Friberg, 2017). Although the proposed method has improved predictions in several aspects relative to previous methods, more improvement is possible and is the subject of further research. Future work related to incorporating measurement error as well as predicting causes of mortality with increased resolution for narrower age groups is ongoing and is likely to increase the usability and reliability of cause-of-death estimates.

## Data Availability

The data used for this work are publicly available on github.

https://github.com/amulick/MCEE-neo.git

## Funding

Bill and Melinda Gates Foundation, grant number OPP1096225

## Conflicts of interest

None declared.

## Notes

### Competing Interest Statement

The authors have declared no competing interest.

### Funding Statement

This work was funded by the Bill and Melinda Gates Foundation, grant number OPP1096225.

### Author Declarations

Data used for this research was collected from previously published papers and publicly available national statistics. No new data was collected.

### Summary of Updates

This version includes more detail on the input studies (new text in sections 1.1 and 3.2 and a new supplemental figure e1), a new description of how the misclassification matrix was specified (new section 3.1), and several corrections and clarifications in the methods, model building process, results and discussion (sections 2-5).

