## Supplementary Material for "A Bayesian hierarchical model with integrated covariate selection and misclassification matrices to estimate neonatal and child causes of death"

### Text E1

#### Covariate selection

The frequentist covariate selection approach used a version of forward stepwise selection where we sequentially added covariates depending on the extent to which they improved the out-of-sample goodness of fit (GOF) under a jackknife (i.e. leave-one-out) procedure. The out-of-sample GOF is calculated for the left-out samples using the chi-squared GOF metric. This covariate selection process uses binomial logistic regressions with the outcome defined as ln(cause/intrapartum). The following steps were performed for each cause equation to select the set of covariates for inclusion in the multinomial regression: 1) perform univariate regressions between the outcome and each covariate and calculate the respective chi-squared statistic, 2) use the best ranked covariate to begin building the multivariable regression, and 3) add to the multivariable regression the covariate with next best GOF which is not already in the regression. If the overall GOF was worse or the same, then the covariate was dropped and the next best fitting covariate was added. If the overall GOF was better, then the covariate was retained and step 3 was repeated with the remaining covariates.

#### Multi-cause multinomial logistic regression

In step 2 we used a multi-cause multinomial logistic regression model – where all CODs are modelled simultaneously – for several reasons. First, the multinomial distribution assigns each death to one cause category, which reflects the way that cause-of-death statistics are generally presented, and ensures that the proportions of deaths attributed to each different cause add to 1. Additionally, if a specific cause is not explicitly reported in a study, the multinomial model can be modified to incorporate the possibility that some deaths due to that cause did occur but were recorded in a different cause category. This is not possible when using some alternative modelling strategies (e.g. a set of binomial models).

The high mortality multinomial model included seven equations to model the eight key neonatal COD categories (i.e. one equation for each non-baseline cause). To account for the potential lack of independence of the assignment of causes of death within a given study, we gave individual deaths from larger studies less weight than those from smaller ones. We did this by giving each death in a particular study a weight inversely proportional to the square root of the total number of deaths contributed by that study. This is intermediate between giving equal weight to each death versus each study in the input data.

### Text E2

#### Model input data

Figure e1. Flow chart illustrating sources of the input data. For calculating cause of death distributions and credible intervals in high-mortality countries without nationally representative input data, random effects were randomly drawn from all studies. In such countries with nationally representative data, random effects were drawn from relevant studies in the green box.

95 studies in the input data

68 from countries considered high-mortality during the prediciton period (2000-2015)

10 nationally representative

58 not nationally representative

27 from countries not considered high-mortality during the prediction period

6 nationally representative

21 not nationally representative

#### Bayesian modelling details

To begin we ran 11 models with values of $\lambda$ ranging from 0.1 (little to no penalisation) to 1000 (very strong penalisation) in increments of 100, fixing $b$ at 0.07, and inspected the constraining behaviour on the fixed-effects parameters. Trace plots and Gelman plots indicated parameter and chain convergence for all models except the relatively unconstrained model where $\lambda$=0.1; the multivariate Gelman statistic also indicates this in **figure e2**. We used trace plots from the $\lambda$=100 model to determine MCMC iterations.

Figure e2. Multivariate Gelman statistic plotted against fixed-effects shrinkage parameter lambda from the Bayesian neonatal cause-of-death model

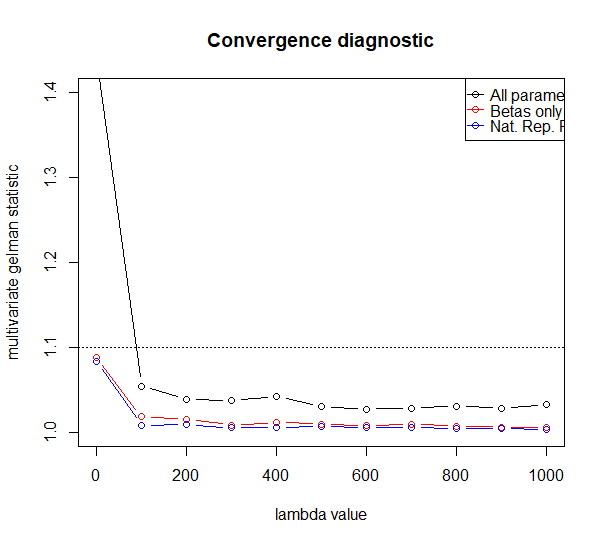

#### Cross-validation

To determine the range of $\lambda$ values in which to narrow our focus for cross-validation, we examined the median values of the $\beta$ coefficients from the posterior distributions. We narrowed our search to $\lambda$s that produced some, but not complete, shrinkage of coefficients. At $\lambda$=300 nearly half the coefficients over all causes were effectively zero (**figure e3**). We discarded $\lambda$>300 and narrowed our search range several times on smaller increments below this until we chose a range of 5 and 10 to 250 in increments of 20.

Figure e3. Median beta coefficients from the posterior distributions of our Bayesian neonatal cause-of-death model plotted against log(lambda), shown together and separately for each COD equation.

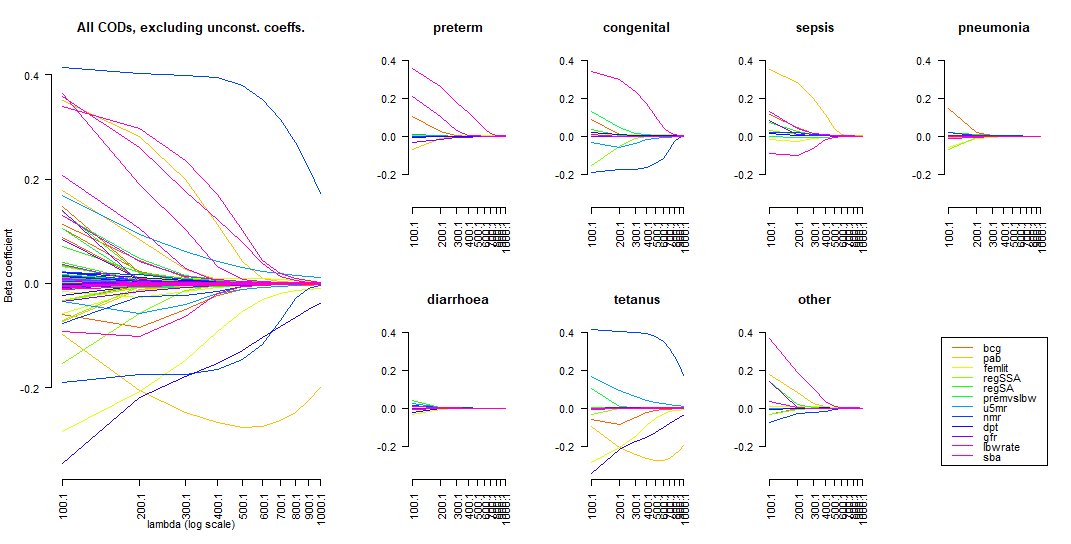

To ensure comparability within and between each combination we used the same starting seed for the random number generator in all 95 x n(lambda) x n(b) computational models.

### Text E3

#### Computational considerations

To minimise run time within a single model, we used the parallel processing R package “doParallel” to distribute non-sequential computing tasks over multiple processors of a high performance computing (HPC) cluster onsite at LSHTM.

We ran the first models on a 2.8 gigaHertz (GHz) machine with four parallel processors. Each 4-chain, 20,000-iteration, eight-outcome multinomial regression was on a dataset of 124 observations on 14 covariates and completed in ~1 hour using approximately 6 gigabytes (GB) of random access memory (RAM).

The cross-validation models were run on a 2.8GHz machine with eight parallel processors. Each model (i.e. one $\lambda$ and one $b$) ran 95 similar regressions and completed in ~16.5 hours using approximately 26GB of RAM. Each of these regressions iterated 10,000 times on datasets containing 121-123 observations and, to reduce memory load, saved only the posterior means, medians and SDs from the simulated data.
